# Adapting the intensity gradient for use across commonly derived accelerometer activity metrics: A LABDA Network project

**DOI:** 10.1101/2025.07.11.25331383

**Authors:** Henrik R. Eckmann, Cameron Razieh, Sebastien Chastin, Lauren B. Sherar, Bjørge H. Hansen, Alex V. Rowlands

**Affiliations:** Diabetes Research Centre, Leicester General Hospital, University of Leicester, UK; National Institute for Health Research (NIHR) Leicester Biomedical Research Centre (BRC), University Hospitals of Leicester NHS Trust and the University of Leicester, Leicester, UK; Leicester Real World Evidence Unit, Diabetes Research Centre, Leicester General Hospital, University of Leicester, Leicester, UK; Office for National Statistics, Newport, NP10 8XG, UK; School of Health and Life Sciences, Glasgow Caledonian University, Glasgow G4 0BA, UK; School of Sport, Health and Exercise Sciences, Loughborough University, Loughborough, UK; Department of Sport Science and Physical Education, University of Agder, Kristiansand, Norway

**Keywords:** physical activity, agreement, intensity distribution

## Abstract

The intensity gradient (IG) quantifies the distribution of time spent across accelerometer-assessed physical activity intensity and is positively associated with health. It was developed using the Euclidean Norm Minus One (ENMO) intensity metric. This study aimed to enable generation of comparable IGs across other metrics (mean amplitude deviation (MAD), monitor independent movement summary units (MIMS), and counts), by addressing a key step in the IG algorithm of dividing physical activity intensity into incremental intensity bins. Two methods of creating analogous bins for MAD, MIMS and counts were explored: 1) linear scaling (“naïve”); 2) non-linear modelling (“modelled”). Generated IGs were compared to the original IG (IG_ENMO) using limits of agreement (LoA) and intra-class correlation (ICC). 43 adults (age, median [IQR]: 23 (21, 26), 61% female) were included. Relative to IG_ENMO, the modelled approach led to lower IGs (bias: −0.43, −1.23, −0.91 for MAD, MIMS, and counts, respectively). In contrast, the naïve approach led to higher IGs (+0.27, +0.39, +0.54, respectively). For MAD and counts, LoA were slightly wider for naïve bins (95% LoA: ±0.26, ±0.34) vs modelled bins (±0.21, ±0.28), but for MIMS were slightly wider for modelled bins (modelled: ±0.35, naïve: ±0.31). ICCs were higher for the modelled approach with IG_MAD most consistent (ICC 95% confidence interval: 0.72-0.91) and IG_MIMS least consistent (0.59-0.86). For the naïve approach, IG_MAD was most consistent (0.49-0.82) and IG_counts least consistent (0.09-0.61). Results indicate that consistency of the IG between metrics is improved with appropriate scaling to create analogous intensity bins, but agreement is limited.

## Introduction

The intensity gradient (IG) is a summary measure of the distribution of the intensity of accelerometer-assessed daily physical activity. Since its introduction in 2018 (Rowlands et al., 2018), use of the IG in studies of cardiometabolic risk(Dawkins, Yates, Edwardson, et al., 2022), waist-to-height-ratio and metabolic syndrome risk(Fairclough et al., 2019), incident cardiovascular disease (Dempsey et al., 2022), life expectancy(Zaccardi et al., 2024), and all-cause mortality(Schwendinger et al., 2024) has improved our understanding of the role of the intensity distribution, independent of and in addition to the volume, of physical activity. The IG can be generated in the widely used accelerometer processing R package GGIR(Migueles et al., 2019), and has been used in a variety of studies, usually with the Euclidean Norm Minus One (ENMO)(van Hees et al., 2013) as the epoch-level intensity metric as in the original implementation(Dawkins, Yates, Edwardson, et al., 2022; Dawkins, Yates, Razieh, et al., 2022; Dempsey et al., 2022; Fairclough et al., 2019; Hernández-Vicente et al., 2022; Maylor et al., 2023), but sometimes applied to other epoch-level metrics in use in accelerometer research, e.g. ActiGraph counts(Aadland et al., 2021; Willems et al., 2024), and Mean Amplitude Deviation (MAD)(Edwardson et al., 2022; Willems et al., 2024).

With increasing implementation of the IG in research, there is a clear need to assess the impacts of applying it to epoch-level metrics other than ENMO, and how best to apply it, including potential necessary modifications to the algorithm. To understand why the IG may need modification based on the epoch-level metric it is applied to, we must first consider the IG algorithm. The original implementation of the IG has previously been described in detail (Rowlands et al., 2018). In brief, each 5-second ENMO epoch is counted into incremental bins of 25 (0-25, 25-50, 50-75, …, 3975-4000, >4000) milli-gravitational units (m*g*). Plotting a histogram of the number of epochs in each bin shows the distribution of measured intensity at the epoch level for a given day, typically displaying a characteristic negative curvilinear relationship between the intensity and number of observations, reflecting the fact that most people spend most of their day at low intensities of movement and progressively less time at higher intensities. Assigning each bin the value at its midpoint (e.g. 0-25 becomes 12.5 m*g*) and log transforming both axes produces an approximately linear relationship between intensity and number of observations, indicating that the relationship follows a power law distribution(Pinto et al., 2012). The IG is the estimated slope of the relationship, equivalent to the exponent of the underlying power law distribution and thus describes the relationship between time spent at high versus low intensities. A steeper slope, indicated by a lower/more negative value, indicates a higher ratio of low intensity observations and a shallower slope indicates a more even distribution of time spent across intensities.

The mapping of each 5-second epoch into 25 m*g* bins is of particular interest because bin values will impact the estimation of the IG and potentially the association with health. For MAD, which is also measured in m*g*, using the same values as ENMO for bins may seem logical, but for other epoch-level metrics which are measured in other, sometimes arbitrary, units on very different scales (e.g. counts and MIMS), no obvious equivalent bins exist. We hypothesised that the bin values for a given epoch-level metric would at least need to be of a similar scale compared to the expected range of values for that epoch-level metric as 25m*g* is to the expected range of values for ENMO.

Recently, studies examining the impact of applying the IG to MAD, counts, and raw vector magnitude without modification have been published(Alexander et al., 2024; Clevenger et al., 2025; Willems et al., 2024) showing overall limited agreement between epoch-level metrics. Two of these studies further proposed simple modifications for counts(Alexander et al., 2024; Clevenger et al., 2025), MAD, and raw vector magnitude(Clevenger et al., 2025), without major improvements for agreement. The logic behind the modifications suggested by Clevenger et al. aligns with our hypothesis, but the lack of improvement found suggests that simple linear scaling is not sufficient. This is supported by a study by Karas et al. demonstrating that mapping between epoch-level metrics depends on the specifics of each metric and cannot be assumed to be linear(Karas et al., 2022). Thus, to investigate the benefit of a more bespoke approach, we assessed bins generated through both linear scaling (“naïve bins”) and non-linear modelling (“modelled bins”). We chose to include the Monitor Independent Movement Summary (MIMS) rather than raw vector magnitude, as it is the epoch-level metric that has been used with the US National Health and Nutrition Examination Survey (NHANES) data(Belcher et al., 2021), and so far has not been used with the IG.

The aim of this study was to adapt the IG for use across other accelerometer metrics by 1) generating appropriate IG bins for each of MAD, MIMS, and counts, and 2) assessing the agreement between the IGs generated using different epoch-level intensity metrics.

## Materials and Methods

### Study design

To address the aims of this study we generated ENMO, MAD, MIMS, and counts from a single accelerometer device for each participant. From these data, two candidate sets of analogous bins for each of MAD, MIMS, and counts were derived using a simple linear approach (“naive bins”) and a non-linear approach specific to the association between ENMO and the alternate epoch-level metric (“modelled bins”). The IG was then calculated for each combination of epoch metric and candidate set of bins, and the agreement between each of them and IG derived from EMNO assessed. Each step is described in detail in the following sections.

### Study population

We conducted a secondary analysis of data collected in a sample of 56 adult participants from the University of Leicester and Loughborough University, UK collected between November 2017 to July 2018 and previously described in detail(Rowlands et al., 2019). All participants provided written informed consent, and the study was approved by ethics representatives from the College of Life Sciences, University of Leicester. Participants were asked to wear multiple accelerometers, but for this analysis data from only one accelerometer was used, the GENEActiv (Activinsights Ltd, Cambridgeshire, UK). This was worn on the non-dominant wrist 24 hours a day for four days, recording at a sample rate of 100 Hz and a dynamic range of +/-8 gravitational units (*g*).

### Data Processing

To generate all four metrics while benefitting from other functionality built into the commonly used R package GGIR(Migueles et al., 2019), a custom processing pipeline was built recreating relevant steps in GGIR or calling specific GGIR functions directly where possible, bypassing the usual recommended GGIR wrapper function. Thus, the data were processed equivalently to using GGIR (parts 1 and 2) in the standard way, except with MIMS generated alongside ENMO, MAD, and counts in 5-second epochs. This included auto-calibration(van Hees et al., 2014), clipping-, and non-wear detection using the current standard GGIR algorithms. Participants were excluded if calibration failed using the calibration error threshold of 0.01 *g*(Rowlands et al., 2018).

To address our first aim, analogous bins for MAD, MIMS, and counts were generated, which is described in the statistical analysis section. Following this, to address our second aim, epoch-level data were fed back into GGIR to generate the mean daily average acceleration(Rowlands, 2018) (AvAcc) and IG for each participant for each epoch-level metric using default GGIR settings, except for the metric-specific IG which was generated using the metric-specific bins generated in our first aim (see statistical analysis section).

Mean daily AvAcc and IG for each participant were calculated excluding participants with fewer than three valid wear days to align our protocol with the one used in the original IG paper(Rowlands et al., 2018). As in that paper, a valid day was defined as any day with at least 16 hours of valid data and for which the participant had data covering the whole 24 hours across the measurement period to allow imputation of non-wear and clipped data. As a result, more participants were analysed for the first aim (generation of appropriate bins) than the second aim (assessing the agreement of the IGs generated using different epoch-level metrics).

### Accelerometer epoch-level metrics

ENMO is the epoch mean of the vector magnitude (using the Euclidean norm) of the triaxial acceleration signal, minus 1 *g* to account for the influence of gravity, and is thus a measure of the mean magnitude of acceleration within the epoch(van Hees et al., 2013).

MAD is also based on the vector magnitude (using the Euclidean norm) but measures the mean absolute deviation from the epoch mean, rather than simply the epoch mean directly(Vähä-Ypyä et al., 2015). MAD is thus a measure of variation rather than absolute magnitude and as such, unlike ENMO, does not need to account for the influence of gravity as gravity is static and variation in the signal therefore must come from movement.

Counts are derived from a comparatively more complex algorithm consisting of multiple resampling and filtering (meant to account for gravity and signal noise) steps for each axis of the raw acceleration signal. Each axis is rectified, capped at a max value of 128 (due to signed 8-bit integers having a range of −128 to 127 and the early ActiGraph devices using 8-bit memory(Tryon & Williams, 1996)), and then summed over the epoch(Neishabouri et al., 2022). Triaxial counts are calculated by taking the vector magnitude (using the Euclidean norm) of the counts from each individual axis and are a measure of the magnitude of acceleration in the epoch, like ENMO.

The MIMS algorithm attempts to overcome differences in specifications between devices by interpolating each axis to a sampling frequency of 100 Hz and extrapolating in areas where clipping is detected due to limited dynamic range. The signal is then filtered to account for gravity and signal noise, like counts, and the area under the curve of the rectified signal is calculated for each epoch. Rather than using the Euclidean norm, triaxial MIMS are based on the vector magnitude using the Manhattan norm(John et al., 2019). MIMS is thus a measure of vector magnitude, like ENMO and counts, although using a different norm.

Because ENMO and MAD are each based on means over the epoch while counts and MIMS are based on sums, for a given sustained acceleration, the magnitude of counts and MIMS will scale with epoch length, while the magnitude of ENMO and MAD will not.

### Statistical Analyses

Descriptive data were reported as median (interquartile range) or count (percentage).

#### Aim 1: Generation of IG bins for MAD, MIMS, and counts

As indicated in the introduction of this paper, two approaches to generating analogous bins for each of MAD, MIMS, and counts were explored: 1) simple linear scaling of the ENMO bins (“naïve bins”) and 2) modelling ENMO into each metric and predicting equivalent values for each bin (“modelled bins”). For this step, all valid epochs were used, meaning non-wear and clipped data were removed, no imputation was undertaken, and no restrictions on amounts of valid wear per participant or day were imposed.

For the naïve bins, the 25 m*g* bins were scaled by the ratio of the left-most edge of the top bin (4000 m*g*) to the value at the corresponding percentile for each epoch-level metric. Note that this method is similar to the one used by Clevenger et al.(Clevenger et al., 2025)

For the modelled bins, multiple competing linear regression models were compared, including 1) a model fitting ENMO as a linear variable, and 2) five models fitting ENMO as a non-linear variable using restricted cubic splines with progressively more knots (3 to 7) (Harrell, 2015). Due to highly positively skewed data, placing knots at standard percentiles was ineffective. Instead, *n* knots were placed at the 0^th^ through the *n*-1^st^ power of the *n*-1^st^ root of the left edge of the topmost bin, i.e. 4000 ENMO. This way the outer knots were 1 and 4000 for all models, and knots in between were distributed across the range but more concentrated at the lower end. The best model was selected based on the Bayesian Information Criterion (BIC) (i.e. lowest BIC)(Schwarz, 1978) for each epoch-level metric. For all epoch-level metrics, the lowest BIC (best model) was found when including non-linear associations (**Supplementary Table S1**). Each chosen model was then used to predict equivalent values for each bin edge, except for the lower edge of the lowest bin and the higher edge of the highest bin. The lower edge of the lowest bin was set at 0 for all epoch-level metrics and the higher edge of the highest bin was set to be double the value of the lower edge, as it is for ENMO, to avoid data-points above the highest bin.

For each approach, the agreement of each of MAD, MIMS, and counts with ENMO in terms of classifying each data-point into corresponding bins was assessed by median- and mean difference, and weighted Kappa(Cohen, 1968).

#### Aim 2: Agreement between the IGs generated using different epoch-level metrics

Intraclass Correlation Coefficients(Koo & Li, 2016) (ICC, two-way mixed effects model, consistency, single rater), mean bias, and limits of agreement (LoA)(Bland & Altman, 1986) between IG generated from ENMO and IG generated from each of MAD, MIMS, and counts using either the modelled or naïve bins were calculated. ICC model was selected and reliability categorised in accordance with Koo and Li, 2016(Koo & Li, 2016). Presented values for the naïve bins were rounded to the nearest m*g*, count, and thousandth of a MIMS unit respectively. Analyses of agreement between rounded and non-rounded values were carried out to assess the impact of rounding.

This analysis was completed using statistical software R version 4.3.1.

## Results

### Aim 1: Generation of IG bins for MAD, MIMS, and counts

Of the 56 participants in the cohort, data from 54 (median [interquartile range] age: 24 [21, 27] years; height: 171.3 [163.3, 180] cm; mass: 63.4 [57, 80] kg; 65% female) were used for modelling (**Table 1**). One participant was excluded during data collection for technical reasons, and one was excluded during processing due to a technical error when trying to extract data from the accelerometer data-file (**Supplementary Figure S1**).

**Table 1.**
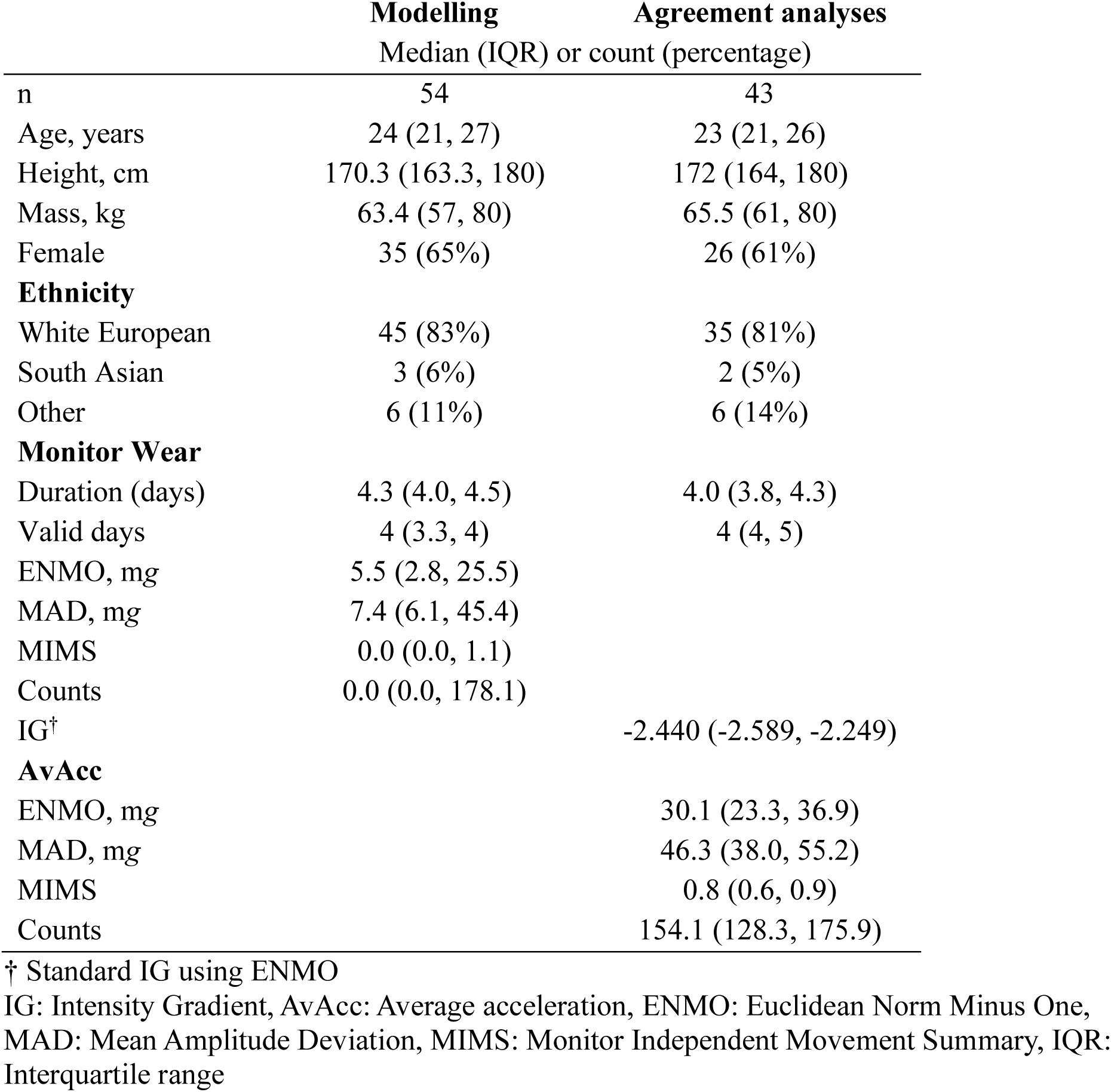
Descriptive statistics.

|  | **Modelling** | **Agreement analyses** |
| --- | --- | --- |
|  | Median (IQR) or count (percentage) |  |
| n | 54 | 43 |
| Age, years | 24 (21, 27) | 23 (21, 26) |
| Height, cm | 170.3 (163.3, 180) | 172 (164, 180) |
| Mass, kg | 63.4 (57, 80) | 65.5 (61, 80) |
| Female | 35 (65%) | 26 (61%) |
| **Ethnicity** |  |  |
| White European | 45 (83%) | 35 (81%) |
| South Asian | 3 (6%) | 2 (5%) |
| Other | 6 (11%) | 6 (14%) |
| **Monitor Wear** |  |  |
| Duration (days) | 4.3 (4.0, 4.5) | 4.0 (3.8, 4.3) |
| Valid days | 4 (3.3, 4) | 4 (4, 5) |
| ENMO, mg | 5.5 (2.8, 25.5) |  |
| MAD, mg | 7.4 (6.1, 45.4) |  |
| MIMS | 0.0 (0.0, 1.1) |  |
| Counts | 0.0 (0.0, 178.1) |  |
| IG<sup>†</sup> |  | -2.440 (-2.589, -2.249) |
| **AvAcc** |  |  |
| ENMO, mg |  | 30.1 (23.3, 36.9) |
| MAD, mg |  | 46.3 (38.0, 55.2) |
| MIMS |  | 0.8 (0.6, 0.9) |
| Counts |  | 154.1 (128.3, 175.9) |

<sup>†</sup> Standard IG using ENMO
IG: Intensity Gradient, AvAcc: Average acceleration, ENMO: Euclidean Norm Minus One, MAD: Mean Amplitude Deviation, MIMS: Monitor Independent Movement Summary, IQR: Interquartile range

Of the candidate models, the lowest BIC was achieved fitting ENMO as a non-linear variable, with splines of 7 knots for MAD and MIMS, and 6 knots for counts, with R-squared values of 0.96, 0.88, and 0.81, respectively (**Figure 1**).

**Figure 1.**
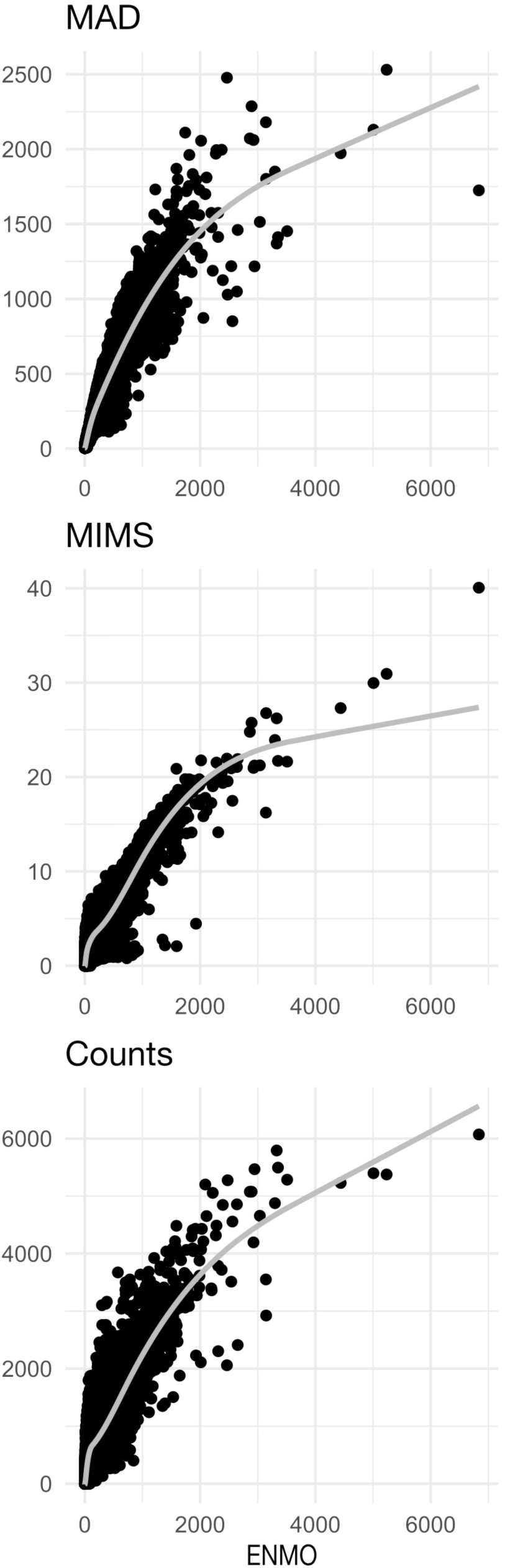
Scatterplots with model-fit overlaid for chosen models. ENMO on the x-axes and each of MAD (top), MIMS (middle), and counts (bottom) on the y-axes. Points represent a subset of the data created by taking every 10^th^ observation. MAD: Mean Amplitude Deviation, MIMS: Monitor Independent Movement Summary, ENMO: Euclidean Norm Minus One

Naïve bins after rounding were 15 m*g*, 0.173 MIMS, 34 counts for MAD, MIMS, and counts respectively. Agreement analyses to assess the impact of rounding on IG results showed no significant impacts. Results are presented in **Supplementary Table S2**. Due to the non-linear nature of the modelling underlying the modelled bins, the spacing of bin cut points is progressively smaller as the intensity increases. The modelled bins are included in **Supplementary Table S4.**

Classification accuracy (weighted Kappa, mean-, and median difference) is shown in **Table 2**. MAD showed the highest agreement compared with ENMO for both the naïve (Kappa: 0.72) and modelled (0.95) bin approaches, MIMS the second highest (naïve bins: 0.52; modelled bins: 0.90), and counts the lowest (naïve bins: 0.46; modelled bins: 0.81). Agreement was higher for the modelled approach vs. the naïve for each metric. For MAD, MIMS and counts, the naïve approach showed a tendency (**Figure 2a**) for classifying each point in a higher bin compared with ENMO, most pronounced towards the middle of the range. In contrast, the modelled approach showed errors spread more evenly in both directions (**Figure 2b**).

**Figure 2.**
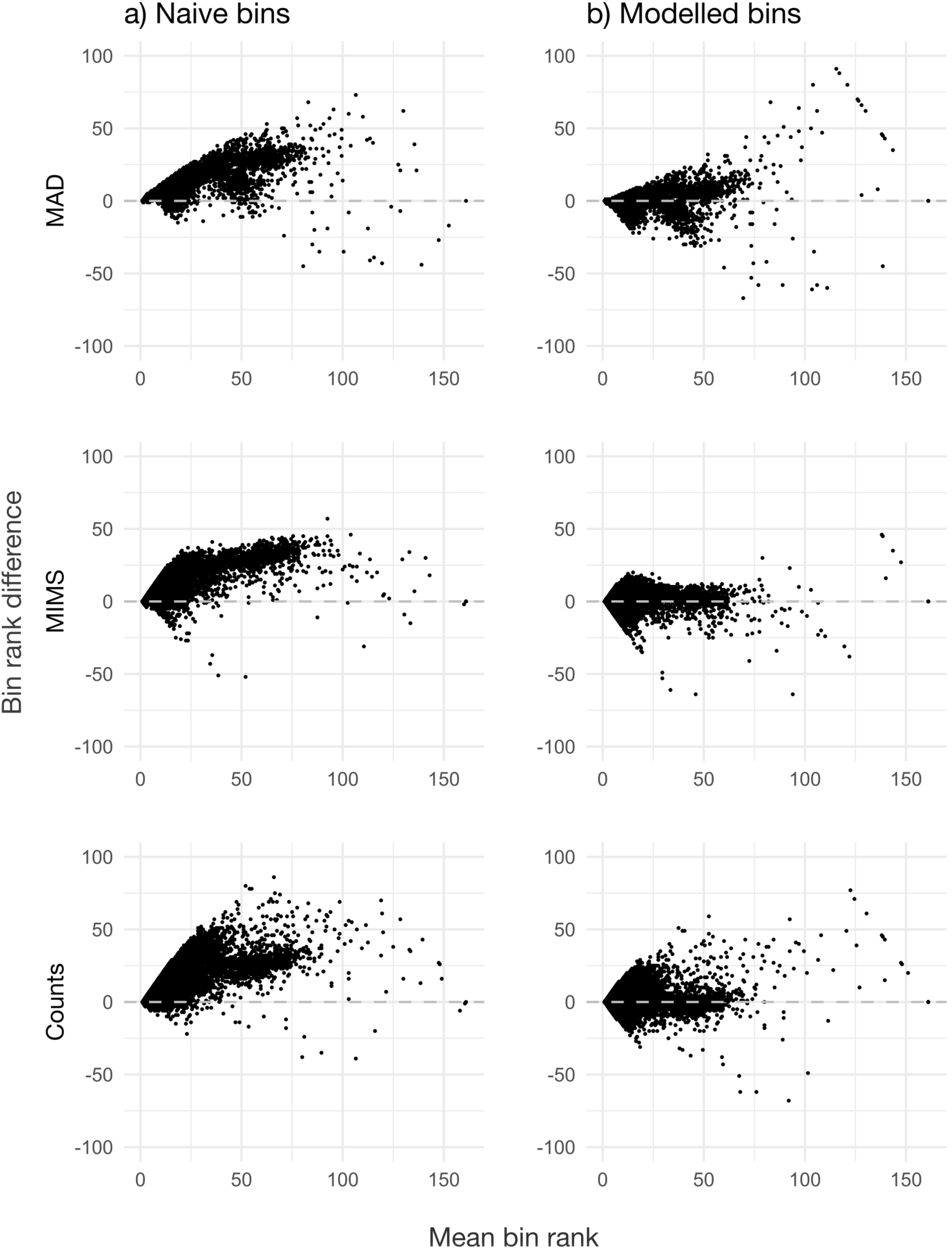
Classification accuracy for naïve approach (a) and modelled approach (b). Scatterplots with mean bin rank for paired observations on the x-axes and the differences (each of MAD (top), MIMS (middle), and counts (bottom) minus ENMO) between the same on the y-axes. Points represent a subset of the data created by taking every 10^th^ observation. MAD: Mean Amplitude Deviation, MIMS: Monitor Independent Movement Summary, ENMO: Euclidean Norm Minus One

**Table 2.** Classification accuracy metrics.

|  | **Kappa<sup>†</sup>** | **Mean** | **Median** |
| --- | --- | --- | --- |
| **Naïve bins** |  |  |  |
| MAD | 0.72 | 2.79 | 1 |
| MIMS | 0.52 | 3.53 | 1 |
| Counts | 0.46 | 3.57 | 1 |
| **Modelled bins** |  |  |  |
| MAD | 0.95 | 1.95 | 1 |
| MIMS | 0.90 | 2.00 | 1 |
| Counts | 0.81 | 2.05 | 1 |

<sup>†</sup> 95% CIs all +/- <0.004
Weighted Kappa, mean- and median bin rank difference for data points classified based on each of MAD, MIMS, and counts with bins according to the naïve or the modelled approach minus ENMO with 25 mg bins.
MAD: Mean Amplitude Deviation, MIMS: Monitor Independent Movement Summary, ENMO: Euclidean Norm Minus One

### Aim 2: Agreement between the IGs generated using different epoch-level metrics

For the IG agreement analysis, a further 11 participants were excluded due to insufficient valid wear leading to a sample of 43 (median [IQR] age: 23 [21, 26] years; height: 172 [164, 180] cm; mass: 65.5 [61, 80] kg; 61% female) participants.

Mean biases (each of MAD, MIMS, and counts minus ENMO) were positive for each epoch-level metric for the naïve approach, with MAD showing the least bias, and counts showing the highest (**Table 3**). Conversely, mean biases were negative for the modelled approach with MAD again showing the least bias but with MIMS showing the highest. The magnitude of the mean bias tended to be higher for the modelled approach compared with the naïve (**Figure 3**), while LoA tended to be slightly narrower for MAD and counts, but slightly wider for MIMS. Modelled MIMS, and all three naïve metrics showed a proportional bias, whereby bias decreased as intensity increased.

**Figure 3:**
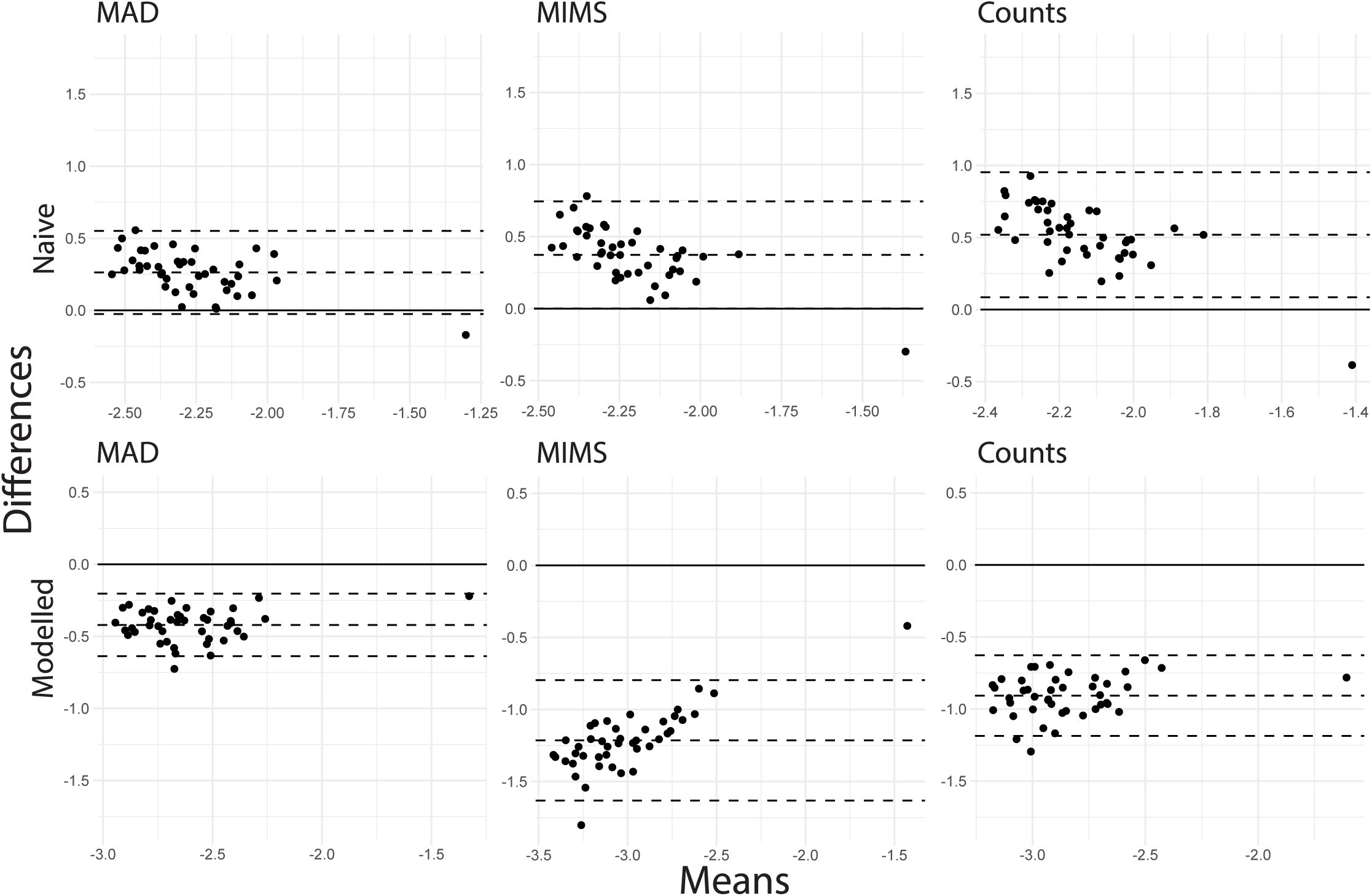
Bland-Altman plots showing bias and limits of agreement for intensity gradients generated based on naïve and modelled approaches for each of MAD, MIMS, and counts compared to ENMO. X-axes: means of paired IGs, y-axes: differences between paired IGs (each of MAD, MIMS, and counts minus ENMO). IG: Intensity gradient, MAD: Mean Amplitude Deviation, MIMS: Monitor Independent Movement Summary, ENMO: Euclidean Norm Minus One

**Table 3:** Agreement analysis metrics.

|  | **Bias<sup>†</sup>** | **LoA<sup>†</sup>** | **ICC (95% CI)** | **IG (median (IQR))** |
| --- | --- | --- | --- | --- |
| ENMO |  |  |  | -2.440 (-2.589, -2.249) |
| **Naïve bins** |  |  |  |  |
| MAD | 0.26 | -0.03; 0.55 | 0.79 (0.64; 0.88) | -2.174 (-2.260, -2.052) |
| MIMS | 0.37 | 0.00; 0.74 | 0.59 (0.36; 0.76) | -2.063 (-2.110, -1.949) |
| Counts | 0.52 | 0.01; 0.95 | 0.41 (0.13; 0.63) | -1.886 (-1.937, -1.815) |
| **Modelled bins** |  |  |  |  |
| MAD | -0.42 | -0.64; -0.20 | 0.92 (0.86; 0.96) | -2.835 (-2.988, -2.713) |
| MIMS | -1.21 | -1.64; -0.80 | 0.82 (0.69; 0.90) | -3.685 (-3.905, -3.427) |
| Counts | -0.91 | -1.19; -0.63 | 0.87 (0.78; 0.93) | -3.358 (-3.500, -3.152) |

<sup>†</sup> Calculated from MAD, MIMS, and counts minus ENMO
Intra-class correlation coefficients, bias and limits of agreement for intensity gradients based on naïve and modelled approaches for each of MAD, MIMS, and counts compared to ENMO.
IG: Intensity gradient, LOA: limits of agreement, ICC: Intra-class correlation coefficient, IQR: Interquartile range, CI: Confidence interval, MAD: Mean Amplitude Deviation, MIMS: Monitor Independent Movement Summary, ENMO: Euclidean Norm Minus One

ICCs were higher for the modelled (0.82-0.92) than the naïve (0.41-0.79) approach, irrespective of metric, and for MAD (0.92 (good to excellent) for modelled approach, 0.79 (good) for naïve approach), irrespective of approach (**Table 3**). MIMS had the lowest ICC (moderate to excellent), with counts in between (good to excellent) for the modelled approach; conversely counts had the lowest ICC for the naïve approach (poor to moderate for counts, poor to good for MIMS).

One outlier, with a very high IG, was apparent from visual inspection of Bland-Altman plots. A separate analysis was conducted excluding the outlier, showing slightly worse ICCs overall but very similar biases and LoA. Results are reported in **Supplementary Table S3** and **Supplementary Figure S2**.

## Discussion

This study adapted the IG for use with epoch-level metrics other than ENMO. To the authors’ knowledge this has only been done in two previous studies; one which investigated using fewer, wider bins for counts only(Alexander et al., 2024), and one which investigated linear scaling of bins for MAD, counts and raw vector magnitude(Clevenger et al., 2025). A further study did compare the IG between ENMO, MAD, and two versions of counts, but without any adaptations to account for the impact of the differing distributions evident across metrics on the resulting IG (Willems et al., 2024).

We tested a linear scaling approach to generating appropriate bins for calculation of the IG based on MAD, MIMS, and counts, along with a more complex non-linear modelling approach, and assessed agreement with the original (ENMO) IG for each. While the modelled approach produced more consistent results, particularly based on ICCs, both approaches generated IGs with limited agreement to the original version. Overall, the magnitude of the IG was not comparable between epoch-level metrics, while the relative agreement, i.e. consistency of the pattern of the IG, was good to excellent for MAD, but lower for MIMS and counts, with the IG from the naïve approach for counts and either approach for MIMS showing clear signs of proportional bias, and wide LoA across the board.

These results are similar to the results from Willems et al.(Willems et al., 2024), but with less proportionality to the biases compared to Willems et al.’s application of the ENMO IG method to MAD and counts, which was applied without adaptation. This suggests that, as with measures of overall activity (e.g. AvAcc, counts per day), the magnitude of the IG should not be compared between epoch-level metrics without careful consideration.

However, adapting the bins used to generate the IG might mitigate the issue somewhat, particularly relating to the proportionality of the bias.

Compared to the linearly scaled bins of (11m*g* for MAD, 34 counts for counts)(Clevenger et al., 2025) from Clevenger et al., the naïve bins in the present study were similar for MAD (15 m*g*) and identical for counts (34 counts) reflecting the similar approach used to derive them. The bias found in the present study for counts with naïve bins was also very similar to that shown by Clevenger et al. for counts with linear scaling, while the bias for MAD herein was halved. Clevenger et al. does not report LoAs or ICCs. As is the case for the present study, Clevenger et al. indicates that linear scaling of bins alone does not sufficiently account for epoch-level metric differences to lead to directly comparable IG outcomes. The more consistent results we found for the IG generated using modelled bins supports the suggestion that methods other than simple linear scaling for generation of analogous bins may be needed (Clevenger et al., 2025).

Alexander et al.(Alexander et al., 2024) also recently investigated generating the IG from counts; they summarised counts in 15-second epochs, in contrast to this study, and the original IG implementation, which used 5-second epochs. Two approaches were tested: 1) “countIG” using bin increments of 25 counts to match the 25 m*g* increments used for the original ENMO IG(Rowlands et al., 2018), and 2) “adjIG” where the authors attempted to match the number of bins containing data to the ENMO IG by using bin increments of 100 counts. The latter approach resulted in a total of 41 bins as opposed to 161 bins in the original IG. This leads to much lower resolution in terms of activity intensity and a bottom bin containing both inactive time and light physical activity. This could change the impact of inactive time on the IG and thus associations with health but remains to be investigated.

Alexander et al.(Alexander et al., 2024) reported that “adjIG” is comparable to the ENMO IG, despite reporting ICC’s (95% CI) of 0.00 (−0.05; 0.12) and 0.27 (0.14; 0.64) for “countIG” and “adjIG” respectively, compared to the ENMO IG, with biases and LoA of −0.712 (−1.293; −0.131) and −0.154 (−0.648, 0.339). By comparison either of the approaches in this study would be considered comparable. However, interpreting the ICCs in line with guidelines from Koo & Li(Koo & Li, 2016), the ICCs reported are indicative of poor agreement, and the LoA ranges are almost four times the standard deviation of the IG from ENMO in the study sample. In this study, while ICCs were generally much higher than in Alexander et al., overall agreement was still limited, though MAD, particularly with the modelled approach, performed reasonably well. It is perhaps unsurprising that MAD produced the IG that was most comparable to ENMO, due to the relative similarity of MAD to ENMO (based on vector magnitude and means). In contrast, MIMS and counts are generated through multiple steps and based on integration and summing, respectively, instead of means. The overall results are in that respect as expected.

Moving forward with adapting the IG for accelerometer metrics other than ENMO, it is worth considering the trade-offs for each approach. The modelled approach in this study arose from a pragmatic desire to be able to generate equivalent IGs irrespective of epoch-level metric.

However, there’s a trade-off between agreement and interpretability, as non-linearly modelling bins equivalent to ENMO for another metric adds an extra layer of complexity to an already somewhat complex interpretation. Conversely, the naïve approach in this study, like the linear scaling in Clevenger et al., is simply a pragmatic approach to dividing the relevant intensity range into bins with a similar resolution to that of the standard IG, avoiding complicating the interpretation. While the overall agreement is still limited using the modelled bins, the improvements in terms of relative agreement, indicated by higher ICCs, could lead to more consistent results for associations of the IG to health outcomes, which would make the trade-off more favourable for the modelled approach.

The main strengths of this study were the comparison of the IG between widely used epoch-level metrics derived in a standardized way from accelerometry measured following a widely used protocol and research-grade accelerometer, all in accordance with the original IG implementation. Notable limitations of this study include, first, the relatively small sample size and the inherent distribution of the free-living accelerometer data. While the sample included generally healthy and active people with free-living data covering a wide range of intensities, free-living accelerometer data is naturally very skewed, becoming very sparse at higher intensities. This complicated the modelling process, necessitating non-standard knot placements, and led to increasing uncertainty at higher intensities. Second, there was a limited number of epoch-level metrics included. One could likely make convincing arguments to include any number of competing metrics. The ones included here were chosen because of their widespread use and thus potential to optimise utility of the results. Similar considerations apply to exclusion of other device wear sites. With the IG originally having been developed using the wrist only however, comparing between epoch-level metrics at the wrist was deemed the obvious first step.

In conclusion, our findings demonstrate that, as is the case for proxy measures of activity volume or overall activity, the magnitude of the IG between accelerometer epoch-level metrics is not directly comparable. This is perhaps not surprising given that mapping between epoch-level metrics depends on the specifics of each metric(Karas et al., 2022), and similar findings from recent studies(Alexander et al., 2024; Clevenger et al., 2025; Willems et al., 2024). However, the relative agreement of the IG between ENMO and the other metrics was more similar, particularly for the MAD metric and for non-linearly modelled bins. It remains to be determined the extent to which the differences observed impact on the ability to detect associations with health and differences between population groups. This is an important next step as each of these epoch-level metrics are being used in studies internationally, including large national cohorts such as NHANES and others. Establishing which of the metrics are better suited to capturing associations between the intensity distribution of physical activity and health could enable a more informed approach to selection of appropriate epoch-level metrics moving forward.

**Supplementary materials:** The following supplementary materials are available: **Supplementary Figure S1**: Exclusion flowchart. **Supplementary Table S1**: Bayesian Information Criterions for each model modelling ENMO to each of MAD, MIMS, and counts. **Supplementary Table S2**: Agreement analyses comparing rounded to non-rounded bins. **Supplementary Table S3:** Agreement analyses for IGs based on naïve and modelled approaches for each of MAD, MIMS, and counts compared to ENMO, with outlier excluded. **Supplementary Figure S2**: Bland-Altman plots showing bias and limits of agreement for intensity gradients generated based on naïve and modelled approaches for each of MAD, MIMS, and counts compared to ENMO with outlier excluded. **Supplementary Table S4**: Generated bins.

## Data Availability

The data that support the findings of this study are available from the corresponding author upon reasonable request.

## Acknowledgements

We would like to acknowledge Charlotte Edwardson for her role in generating this dataset and Elysa Ioannou, Francesca Denton and Shinelle Baptiste for assisting with the original data collection and to thank all participants who volunteered to take part in this study.

This is a summary of independent research funded by UK Research and Innovation (UKRI). All University of Leicester authors are supported by the National Institute for Health and Care Research (NIHR) Leicester Biomedical Research Centre (BRC). C.R. is also supported by Applied Research Collaborations – East Midlands (ARC-EM). The views expressed are those of the author(s) and not necessarily those of UKRI, the NIHR or the Department of Health and Social Care. For the purpose of open access, the author has applied a Creative Commons Attribution license (CC BY) to any Author Accepted Manuscript version arising from this submission.

## Funding

This project is funded by UKRI Guarantee funding (EP/X042464/1) for Horizon Europe MSCA Doctoral Networks HORIZON-MSCA-2021-DN-01 (101072993). The LABDA Network has received funding from the European Union’s Horizon Europe research and innovation program under grant agreement no. 101072993.

## Conflicts of interest

The authors declare that they have no other competing interests.

## Data availability

The data that support the findings of this study are available from the corresponding author upon reasonable request. Statistical analysis code will be made available upon journal publication on GitHub at https://github.com/henrikeckmann

## Author contributions

H.E. and A.R. developed the research question. H.E. and C.R. developed the statistical analysis plan. H.E. undertook the statistical analysis with support from C.R.; A.R. had access to the data. H.E. drafted the manuscript. All authors contributed to the research design and revised the manuscript for important intellectual content. A.R. will act as guarantor for this research.

## Notes

### Competing Interest Statement

The authors have declared no competing interest.

### Author Declarations

We conducted a secondary analysis of data collected in a sample of 56 adult participants from the University of Leicester and Loughborough University, UK collected between November 2017 to July 2018. All participants provided written informed consent, and the study was approved by ethics representatives from the College of Life Sciences, University of Leicester.

## References

Aadland, E., Nilsen, A. K. O., Andersen, L. B., Rowlands, A. V., & Kvalheim, O. M. (2021). A comparison of analytical approaches to investigate associations for accelerometry-derived physical activity spectra with health and developmental outcomes in children. Journal of Sports Sciences, 39(4), 430–438. 10.1080/02640414.2020.1824341

Alexander, C. J., Manske, S. L., Edwards, W. B., & Gabel, L. (2024). Adapting the Intensity Gradient for Use with Count-Based Accelerometry Data in Children and Adolescents. Sensors, 24(10).

Belcher, B. R., Wolff-Hughes, D. L., Dooley, E. E., Staudenmayer, J., Berrigan, D., Eberhardt, M. S., & Troiano, R. P. (2021). US Population-referenced Percentiles for Wrist-Worn Accelerometer-derived Activity. Med Sci Sports Exerc, 53(11), 2455–2464. 10.1249/mss.0000000000002726

Bland, J. M., & Altman, D. G. (1986). Statistical methods for assessing agreement between two methods of clinical measurement. Lancet, 1(8476), 307–310.

Clevenger, K. A., McKee, K. L., E. Elwood, R., Pfeiffer, K. A., & Montoye, A. H. K. (2025). Capturing Free-Living Physical Activity: Impacts of Data Collection and Processing Decisions on Mean Acceleration and Intensity Gradient. Measurement in Physical Education and Exercise Science, 1–11. 10.1080/1091367X.2025.2473904

Cohen, J. (1968). Weighted kappa: nominal scale agreement with provision for scaled disagreement or partial credit. Psychol Bull, 70(4), 213–220. 10.1037/h0026256

Dawkins, N. P., Yates, T., Edwardson, C. L., Maylor, B., Henson, J., Hall, A. P., Davies, M. J., Dunstan, D. W., Highton, P. J., Herring, L. Y., Khunti, K., & Rowlands, A. V. (2022). Importance of Overall Activity and Intensity of Activity for Cardiometabolic Risk in Those with and Without a Chronic Disease. Med Sci Sports Exerc, 54(9), 1582–1590. 10.1249/mss.0000000000002939

Dawkins, N. P., Yates, T., Razieh, C., Edwardson, C. L., Maylor, B., Zaccardi, F., Khunti, K., & Rowlands, A. V. (2022). Differences in Accelerometer-Measured Patterns of Physical Activity and Sleep/Rest Between Ethnic Groups and Age: An Analysis of UK Biobank. J Phys Act Health, 19(1), 37–46. 10.1123/jpah.2021-0334

Dempsey, P. C., Rowlands, A. V., Strain, T., Zaccardi, F., Dawkins, N., Razieh, C., Davies, M. J., Khunti, K. K., Edwardson, C. L., Wijndaele, K., Brage, S., & Yates, T. (2022). Physical activity volume, intensity, and incident cardiovascular disease. Eur Heart J, 43(46), 4789–4800. 10.1093/eurheartj/ehac613

Edwardson, C. L., Maylor, B. D., Dawkins, N. P., Plekhanova, T., & Rowlands, A. V. (2022). Comparability of Postural and Physical Activity Metrics from Different Accelerometer Brands Worn on the Thigh: Data Harmonization Possibilities. Measurement in Physical Education and Exercise Science, 26(1), 39–50. 10.1080/1091367X.2021.1944154

Fairclough, S. J., Taylor, S., Rowlands, A. V., Boddy, L. M., & Noonan, R. J. (2019). Average acceleration and intensity gradient of primary school children and associations with indicators of health and well-being. J Sports Sci, 37(18), 2159–2167. 10.1080/02640414.2019.1624313

Harrell, F. E. (2015). Regression Modeling Strategies: With Applications to Linear Models, Logistic and Ordinal Regression, and Survival Analysis. Springer International Publishing. https://books.google.co.uk/books?id=sQ90rgEACAAJ

Hernández-Vicente, A., Marín-Puyalto, J., Pueyo, E., Vicente-Rodríguez, G., & Garatachea, N. (2022). Physical Activity in Centenarians beyond Cut-Point-Based Accelerometer Metrics. Int J Environ Res Public Health, 19(18). 10.3390/ijerph191811384

John, D., Tang, Q., Albinali, F., & Intille, S. (2019). An Open-Source Monitor-Independent Movement Summary for Accelerometer Data Processing. J Meas Phys Behav, 2(4), 268–281. 10.1123/jmpb.2018-0068

Karas, M., Muschelli, J., Leroux, A., Urbanek, J. K., Wanigatunga, A. A., Bai, J., Crainiceanu, C. M., & Schrack, J. A. (2022). Comparison of Accelerometry-Based Measures of Physical Activity: Retrospective Observational Data Analysis Study. JMIR Mhealth Uhealth, 10(7), e38077. 10.2196/38077

Koo, T. K., & Li, M. Y. (2016). A Guideline of Selecting and Reporting Intraclass Correlation Coefficients for Reliability Research. J Chiropr Med, 15(2), 155–163. 10.1016/j.jcm.2016.02.012

Maylor, B. D., Edwardson, C. L., Clarke-Cornwell, A. M., Davies, M. J., Dawkins, N. P., Dunstan, D. W., Khunti, K., Yates, T., & Rowlands, A. V. (2023). Physical Activity Assessed by Wrist and Thigh Worn Accelerometry and Associations with Cardiometabolic Health. Sensors (Basel), 23(17). 10.3390/s23177353

Migueles, J. H., Rowlands, A. V., Huber, F., Sabia, S., & van Hees, V. T. (2019). GGIR: A Research Community–Driven Open Source R Package for Generating Physical Activity and Sleep Outcomes From Multi-Day Raw Accelerometer Data. Journal for the Measurement of Physical Behaviour, 2(3), 188–196. 10.1123/jmpb.2018-0063

Neishabouri, A., Nguyen, J., Samuelsson, J., Guthrie, T., Biggs, M., Wyatt, J., Cross, D., Karas, M., Migueles, J. H., Khan, S., & Guo, C. C. (2022). Quantification of acceleration as activity counts in ActiGraph wearable. Sci Rep, 12(1), 11958. 10.1038/s41598-022-16003-x

Pinto, C. M. A., Mendes Lopes, A., & Machado, J. A. T. (2012). A review of power laws in real life phenomena. Communications in Nonlinear Science and Numerical Simulation, 17(9), 3558–3578. 10.1016/j.cnsns.2012.01.013

Rowlands, A. V. (2018). Moving Forward With Accelerometer-Assessed Physical Activity: Two Strategies to Ensure Meaningful, Interpretable, and Comparable Measures. Pediatr Exerc Sci, 30(4), 450–456. 10.1123/pes.2018-0201

Rowlands, A. V., Edwardson, C. L., Davies, M. J., Khunti, K., Harrington, D. M., & Yates, T. (2018). Beyond Cut Points: Accelerometer Metrics that Capture the Physical Activity Profile. Med Sci Sports Exerc, 50(6), 1323–1332. 10.1249/mss.0000000000001561

Rowlands, A. V., Plekhanova, T., Yates, T., Mirkes, E. M., Davies, M., Khunti, K., & Edwardson, C. L. (2019). Providing a Basis for Harmonization of Accelerometer-Assessed Physical Activity Outcomes Across Epidemiological Datasets. Journal for the Measurement of Physical Behaviour, 2(3), 131–142. 10.1123/jmpb.2018-0073

Schwarz, G. (1978). Estimating the Dimension of a Model. The Annals of Statistics, 6(2), 461–464. http://www.jstor.org/stable/2958889

Schwendinger, F., Infanger, D., Lichtenstein, E., Hinrichs, T., Knaier, R., Rowlands, A. V., & Schmidt-Trucksäss, A. (2024). Intensity or volume: the role of physical activity in longevity. European Journal of Preventive Cardiology, 32(1), 10–19. 10.1093/eurjpc/zwae295

Tryon, W. W., & Williams, R. (1996). Fully proportional actigraphy: A new instrument. Behavior Research Methods, Instruments, & Computers, 28(3), 392–403. 10.3758/BF03200519

Vähä-Ypyä, H., Vasankari, T., Husu, P., Suni, J., & Sievänen, H. (2015). A universal, accurate intensity-based classification of different physical activities using raw data of accelerometer. Clinical Physiology and Functional Imaging, 35(1), 64–70. 10.1111/cpf.12127

van Hees, V. T., Fang, Z., Langford, J., Assah, F., Mohammad, A., da Silva, I. C., Trenell, M. I., White, T., Wareham, N. J., & Brage, S. (2014). Autocalibration of accelerometer data for free-living physical activity assessment using local gravity and temperature: an evaluation on four continents. J Appl Physiol (1985), *117*(7), 738-744. 10.1152/japplphysiol.00421.2014

van Hees, V. T., Gorzelniak, L., Dean León, E. C., Eder, M., Pias, M., Taherian, S., Ekelund, U., Renström, F., Franks, P. W., Horsch, A., & Brage, S. (2013). Separating movement and gravity components in an acceleration signal and implications for the assessment of human daily physical activity. PLoS One, 8(4), e61691. 10.1371/journal.pone.0061691

Willems, I., Verbestel, V., Dumuid, D., Calders, P., Lapauw, B., & De Craemer, M. (2024). A comparative analysis of 24-hour movement behaviors features using different accelerometer metrics in adults: Implications for guideline compliance and associations with cardiometabolic health. PLoS One, 19(9), e0309931. 10.1371/journal.pone.0309931

Zaccardi, F., Rowlands, A. V., Dempsey, P. C., Razieh, C., Henson, J., Goldney, J., Maylor, B. D., Bhattacharjee, A., Chudasama, Y., Edwardson, C., Laukkanen, J. A., Ekelund, U., Davies, M. J., Khunti, K., & Yates, T. (2024). Interplay between physical activity volume and intensity with modeled life expectancy in women and men: A prospective cohort analysis. Journal of Sport and Health Science, 100970. 10.1016/j.jshs.2024.100970

