## Supplementary materials for "Adapting the intensity gradient for use across commonly derived accelerometer activity metrics: A LABDA Network project"

**Supplementary Table S1: Bayesian Information Criterions for each model modelling ENMO to each of MAD, MIMS, and counts.**

| | | BIC | | |
|---|---|---|---|---|
| | | **MAD** | **MIMS** | **COUNTS** |
| **Linear** | | 37952711.9 | 8423406.9 | 51305973.6 |
| **Non-linear** | | | | |
| Degrees of freedom | **4** | 34837788.8 | 7270772.9 | 50290145.6 |
| | **5** | 33808757.2 | 5586349.1 | 48943938.8 |
| | **6** | 33814358.8 | 5276552.6 | 48975564.3 |
| | **7** | 33819704.5 | 5057159.6 | 48709252.3 |
| | **8** | 33722895.7 | 5027923.4 | 48726050.9 |

BIC: Bayesian Information Criterion, MAD: Mean Amplitude Deviation, MIMS: Monitor Independent Movement Summary, ENMO: Euclidean Norm Minus One

**Supplementary Table S2: Agreement analyses comparing rounded to non-rounded bins.**

| | | | | Rounded | Non-rounded |
|---|---|---|---|---|---|
| | **Bias** | **LoA** | **ICC** (95% CI) | **IG** (median (IQR)) | **IG** (median (IQR)) |
| MAD | 0.01 | -0.04; 0.06 | 0.99 (0.98; 0.99) | -2.173 (-2.265, -2.069) | -2.174 (-2.260, -2.056) |
| MIMS | 0.00 | -0.03; 0.03 | 0.99 (0.99; 0.99) | -2.053 (-2.119, -1.935) | -2.063 (-2.110, -1.960) |
| Counts | 0.00 | -0.00; 0.00 | 0.99 (0.99; 0.99) | -1.886 (-1.937, -1.815) | -1.887 (-1.937, -1.827) |

Intra-class correlation coefficients, bias and limits of agreement for intensity gradients comparing rounded to non-rounded bins for each of MAD, MIMS, and counts.
IG: Intensity gradient, LOA: limits of agreement, ICC: Intra-class correlation coefficient, IQR: Interquartile range, CI: Confidence interval
MAD: Mean Amplitude Deviation, MIMS: Monitor Independent Movement Summary.

**Supplementary Table S3: Agreement analyses for IGs based on naïve and modelled approaches for each of MAD, MIMS, and counts compared to ENMO, with outlier excluded.**

| | **Bias**† | **LoA**† | **ICC** (95% CI) | **IG** (median (IQR)) |
|---|---|---|---|---|
| ENMO | | | | -2.451 (-2.589, -2.253) |
| **Naïve bins** | | | | |
| MAD | 0.27 | -0.01; 0.53 | 0.69 (0.49; 0.82) | -2.174 (-2.260, -2.056) |
| MIMS | 0.39 | 0.08; 0.70 | 0.48 (0.21; 0.68) | -2.063 (-2.110, -1.960) |
| Counts | 0.54 | 0.20; 0.88 | 0.38 (0.09; 0.61) | -1.887 (-1.937, -1.827) |
| **Modelled bins** | | | | |
| MAD | -0.43 | -0.64; -0.22 | 0.84 (0.72; 0.91) | -2.848 (-2.988, -2.717) |
| MIMS | -1.23 | -1.58; -0.89 | 0.76 (0.59; 0.86) | -3.708 (-3.905, -3.471) |
| Counts | -0.91 | -1.19; -0.63 | 0.76 (0.59; 0.86) | -3.359 (-3.500, -3.155) |

† Calculated from MAD, MIMS, and counts minus ENMO

Intra-class correlation coefficients, bias and limits of agreement for intensity gradients based on naïve and modelled approaches for each of MAD, MIMS, and counts compared to ENMO with outlier excluded.

IG: Intensity gradient, LOA: limits of agreement, ICC: Intra-class correlation coefficient, IQR: Interquartile range, CI: Confidence interval

MAD: Mean Amplitude Deviation, MIMS: Monitor Independent Movement Summary, ENMO: Euclidean Norm Minus One

**Supplementary table S4: Generated bins**

| Original bins | Modelled bins | | | Naïve bins | | |
|---|---|---|---|---|---|---|
| **ENMO** | **MAD** | **MIMS** | **Counts** | **MAD** | **MIMS** | **Counts** |
| 0 | 0 | 0 | 0 | 0 | 0 | 0 |
| 25 | 41 | 1.138 | 190 | 15 | 0.173 | 34 |
| 50 | 86 | 1.912 | 399 | 30 | 0.345 | 68 |
| 75 | 128 | 2.351 | 533 | 44 | 0.518 | 101 |
| 100 | 165 | 2.704 | 613 | 59 | 0.690 | 135 |
| 125 | 198 | 2.993 | 658 | 74 | 0.863 | 169 |
| 150 | 228 | 3.231 | 689 | 89 | 1.036 | 203 |
| 175 | 256 | 3.431 | 721 | 104 | 1.208 | 236 |
| 200 | 281 | 3.604 | 755 | 119 | 1.381 | 270 |
| 225 | 305 | 3.763 | 790 | 133 | 1.553 | 304 |
| 250 | 328 | 3.919 | 827 | 148 | 1.726 | 338 |
| 275 | 351 | 4.084 | 866 | 163 | 1.899 | 372 |
| 300 | 373 | 4.259 | 906 | 178 | 2.071 | 405 |
| 325 | 396 | 4.445 | 948 | 193 | 2.244 | 439 |
| 350 | 418 | 4.640 | 991 | 208 | 2.416 | 473 |
| 375 | 440 | 4.844 | 1036 | 222 | 2.589 | 507 |
| 400 | 462 | 5.056 | 1081 | 237 | 2.762 | 540 |
| 425 | 484 | 5.277 | 1127 | 252 | 2.934 | 574 |
| 450 | 505 | 5.505 | 1175 | 267 | 3.107 | 608 |
| 475 | 526 | 5.740 | 1223 | 282 | 3.279 | 642 |
| 500 | 547 | 5.982 | 1271 | 297 | 3.452 | 675 |
| 525 | 568 | 6.230 | 1321 | 311 | 3.625 | 709 |
| 550 | 589 | 6.483 | 1370 | 326 | 3.797 | 743 |
| 575 | 609 | 6.742 | 1420 | 341 | 3.970 | 777 |
| 600 | 629 | 7.005 | 1471 | 356 | 4.142 | 811 |
| 625 | 649 | 7.272 | 1521 | 371 | 4.315 | 844 |
| 650 | 669 | 7.543 | 1572 | 386 | 4.488 | 878 |
| 675 | 689 | 7.817 | 1622 | 400 | 4.660 | 912 |
| 700 | 708 | 8.093 | 1672 | 415 | 4.833 | 946 |
| 725 | 728 | 8.371 | 1722 | 430 | 5.005 | 979 |
| 750 | 747 | 8.651 | 1772 | 445 | 5.178 | 1013 |
| 775 | 765 | 8.932 | 1821 | 460 | 5.351 | 1047 |
| 800 | 784 | 9.214 | 1869 | 475 | 5.523 | 1081 |
| 825 | 802 | 9.496 | 1917 | 489 | 5.696 | 1115 |
| 850 | 821 | 9.777 | 1964 | 504 | 5.868 | 1148 |
| 875 | 839 | 10.058 | 2011 | 519 | 6.041 | 1182 |
| 900 | 857 | 10.337 | 2058 | 534 | 6.214 | 1216 |
| 925 | 874 | 10.614 | 2103 | 549 | 6.386 | 1250 |
| 950 | 891 | 10.889 | 2149 | 564 | 6.559 | 1283 |
| 975 | 909 | 11.160 | 2193 | 578 | 6.731 | 1317 |

| | | | | | | |
|---|---|---|---|---|---|---|
| 1000 | 926 | 11.429 | 2238 | 593 | 6.904 | 1351 |
| 1025 | 942 | 11.693 | 2281 | 608 | 7.077 | 1385 |
| 1050 | 959 | 11.954 | 2325 | 623 | 7.249 | 1419 |
| 1075 | 975 | 12.211 | 2367 | 638 | 7.422 | 1452 |
| 1100 | 991 | 12.464 | 2410 | 653 | 7.594 | 1486 |
| 1125 | 1007 | 12.712 | 2451 | 667 | 7.767 | 1520 |
| 1150 | 1023 | 12.958 | 2492 | 682 | 7.940 | 1554 |
| 1175 | 1039 | 13.199 | 2533 | 697 | 8.112 | 1587 |
| 1200 | 1054 | 13.437 | 2573 | 712 | 8.285 | 1621 |
| 1225 | 1069 | 13.670 | 2613 | 727 | 8.457 | 1655 |
| 1250 | 1084 | 13.901 | 2653 | 742 | 8.630 | 1689 |
| 1275 | 1099 | 14.127 | 2691 | 756 | 8.803 | 1723 |
| 1300 | 1113 | 14.350 | 2730 | 771 | 8.975 | 1756 |
| 1325 | 1128 | 14.569 | 2768 | 786 | 9.148 | 1790 |
| 1350 | 1142 | 14.785 | 2805 | 801 | 9.320 | 1824 |
| 1375 | 1156 | 14.997 | 2842 | 816 | 9.493 | 1858 |
| 1400 | 1170 | 15.206 | 2879 | 831 | 9.666 | 1891 |
| 1425 | 1183 | 15.411 | 2915 | 845 | 9.838 | 1925 |
| 1450 | 1197 | 15.613 | 2951 | 860 | 10.011 | 1959 |
| 1475 | 1210 | 15.812 | 2986 | 875 | 10.183 | 1993 |
| 1500 | 1223 | 16.007 | 3021 | 890 | 10.356 | 2026 |
| 1525 | 1236 | 16.199 | 3055 | 905 | 10.529 | 2060 |
| 1550 | 1249 | 16.387 | 3089 | 919 | 10.701 | 2094 |
| 1575 | 1261 | 16.572 | 3122 | 934 | 10.874 | 2128 |
| 1600 | 1274 | 16.754 | 3156 | 949 | 11.046 | 2162 |
| 1625 | 1286 | 16.933 | 3188 | 964 | 11.219 | 2195 |
| 1650 | 1298 | 17.109 | 3221 | 979 | 11.392 | 2229 |
| 1675 | 1310 | 17.281 | 3253 | 994 | 11.564 | 2263 |
| 1700 | 1322 | 17.451 | 3284 | 1008 | 11.737 | 2297 |
| 1725 | 1333 | 17.617 | 3315 | 1023 | 11.909 | 2330 |
| 1750 | 1345 | 17.781 | 3346 | 1038 | 12.082 | 2364 |
| 1775 | 1356 | 17.941 | 3376 | 1053 | 12.255 | 2398 |
| 1800 | 1367 | 18.099 | 3406 | 1068 | 12.427 | 2432 |
| 1825 | 1378 | 18.253 | 3436 | 1083 | 12.600 | 2466 |
| 1850 | 1389 | 18.405 | 3465 | 1097 | 12.772 | 2499 |
| 1875 | 1400 | 18.554 | 3494 | 1112 | 12.945 | 2533 |
| 1900 | 1410 | 18.700 | 3522 | 1127 | 13.118 | 2567 |
| 1925 | 1420 | 18.843 | 3551 | 1142 | 13.290 | 2601 |
| 1950 | 1431 | 18.983 | 3578 | 1157 | 13.463 | 2634 |
| 1975 | 1441 | 19.121 | 3606 | 1172 | 13.635 | 2668 |
| 2000 | 1451 | 19.256 | 3633 | 1186 | 13.808 | 2702 |
| 2025 | 1460 | 19.388 | 3660 | 1201 | 13.981 | 2736 |
| 2050 | 1470 | 19.518 | 3686 | 1216 | 14.153 | 2770 |

| | | | | | | |
|---|---|---|---|---|---|---|
| 2075 | 1479 | 19.645 | 3712 | 1231 | 14.326 | 2803 |
| 2100 | 1489 | 19.769 | 3738 | 1246 | 14.498 | 2837 |
| 2125 | 1498 | 19.892 | 3763 | 1261 | 14.671 | 2871 |
| 2150 | 1507 | 20.011 | 3788 | 1275 | 14.844 | 2905 |
| 2175 | 1516 | 20.128 | 3813 | 1290 | 15.016 | 2938 |
| 2200 | 1525 | 20.243 | 3838 | 1305 | 15.189 | 2972 |
| 2225 | 1534 | 20.355 | 3862 | 1320 | 15.361 | 3006 |
| 2250 | 1542 | 20.465 | 3886 | 1335 | 15.534 | 3040 |
| 2275 | 1551 | 20.573 | 3909 | 1350 | 15.707 | 3074 |
| 2300 | 1559 | 20.678 | 3933 | 1364 | 15.879 | 3107 |
| 2325 | 1567 | 20.781 | 3956 | 1379 | 16.052 | 3141 |
| 2350 | 1575 | 20.882 | 3978 | 1394 | 16.224 | 3175 |
| 2375 | 1583 | 20.980 | 4001 | 1409 | 16.397 | 3209 |
| 2400 | 1591 | 21.077 | 4023 | 1424 | 16.570 | 3242 |
| 2425 | 1599 | 21.171 | 4045 | 1439 | 16.742 | 3276 |
| 2450 | 1607 | 21.263 | 4066 | 1453 | 16.915 | 3310 |
| 2475 | 1614 | 21.353 | 4088 | 1468 | 17.087 | 3344 |
| 2500 | 1622 | 21.442 | 4109 | 1483 | 17.260 | 3377 |
| 2525 | 1629 | 21.528 | 4129 | 1498 | 17.433 | 3411 |
| 2550 | 1636 | 21.612 | 4150 | 1513 | 17.605 | 3445 |
| 2575 | 1643 | 21.694 | 4170 | 1528 | 17.778 | 3479 |
| 2600 | 1650 | 21.775 | 4190 | 1542 | 17.950 | 3513 |
| 2625 | 1657 | 21.853 | 4210 | 1557 | 18.123 | 3546 |
| 2650 | 1664 | 21.930 | 4230 | 1572 | 18.296 | 3580 |
| 2675 | 1671 | 22.005 | 4249 | 1587 | 18.468 | 3614 |
| 2700 | 1678 | 22.078 | 4268 | 1602 | 18.641 | 3648 |
| 2725 | 1684 | 22.149 | 4287 | 1617 | 18.813 | 3681 |
| 2750 | 1691 | 22.219 | 4306 | 1631 | 18.986 | 3715 |
| 2775 | 1697 | 22.287 | 4325 | 1646 | 19.159 | 3749 |
| 2800 | 1703 | 22.353 | 4343 | 1661 | 19.331 | 3783 |
| 2825 | 1710 | 22.418 | 4361 | 1676 | 19.504 | 3817 |
| 2850 | 1716 | 22.481 | 4379 | 1691 | 19.676 | 3850 |
| 2875 | 1722 | 22.543 | 4396 | 1706 | 19.849 | 3884 |
| 2900 | 1728 | 22.603 | 4414 | 1720 | 20.022 | 3918 |
| 2925 | 1734 | 22.662 | 4431 | 1735 | 20.194 | 3952 |
| 2950 | 1739 | 22.720 | 4448 | 1750 | 20.367 | 3985 |
| 2975 | 1745 | 22.776 | 4465 | 1765 | 20.539 | 4019 |
| 3000 | 1751 | 22.830 | 4482 | 1780 | 20.712 | 4053 |
| 3025 | 1756 | 22.883 | 4499 | 1795 | 20.885 | 4087 |
| 3050 | 1762 | 22.935 | 4515 | 1809 | 21.057 | 4121 |
| 3075 | 1767 | 22.986 | 4531 | 1824 | 21.230 | 4154 |
| 3100 | 1773 | 23.036 | 4547 | 1839 | 21.402 | 4188 |
| 3125 | 1778 | 23.084 | 4563 | 1854 | 21.575 | 4222 |

| | | | | | | |
|---|---|---|---|---|---|---|
| 3150 | 1783 | 23.131 | 4579 | 1869 | 21.748 | 4256 |
| 3175 | 1789 | 23.177 | 4595 | 1883 | 21.920 | 4289 |
| 3200 | 1794 | 23.222 | 4610 | 1898 | 22.093 | 4323 |
| 3225 | 1799 | 23.266 | 4626 | 1913 | 22.265 | 4357 |
| 3250 | 1804 | 23.308 | 4641 | 1928 | 22.438 | 4391 |
| 3275 | 1809 | 23.350 | 4656 | 1943 | 22.611 | 4425 |
| 3300 | 1814 | 23.391 | 4671 | 1958 | 22.783 | 4458 |
| 3325 | 1819 | 23.431 | 4686 | 1972 | 22.956 | 4492 |
| 3350 | 1824 | 23.470 | 4701 | 1987 | 23.128 | 4526 |
| 3375 | 1828 | 23.508 | 4716 | 2002 | 23.301 | 4560 |
| 3400 | 1833 | 23.545 | 4730 | 2017 | 23.474 | 4593 |
| 3425 | 1838 | 23.582 | 4745 | 2032 | 23.646 | 4627 |
| 3450 | 1843 | 23.618 | 4759 | 2047 | 23.819 | 4661 |
| 3475 | 1847 | 23.653 | 4773 | 2061 | 23.991 | 4695 |
| 3500 | 1852 | 23.687 | 4787 | 2076 | 24.164 | 4728 |
| 3525 | 1856 | 23.721 | 4801 | 2091 | 24.336 | 4762 |
| 3550 | 1861 | 23.754 | 4815 | 2106 | 24.509 | 4796 |
| 3575 | 1865 | 23.786 | 4829 | 2121 | 24.682 | 4830 |
| 3600 | 1870 | 23.818 | 4843 | 2136 | 24.854 | 4864 |
| 3625 | 1874 | 23.849 | 4857 | 2150 | 25.027 | 4897 |
| 3650 | 1879 | 23.880 | 4871 | 2165 | 25.199 | 4931 |
| 3675 | 1883 | 23.911 | 4884 | 2180 | 25.372 | 4965 |
| 3700 | 1888 | 23.941 | 4898 | 2195 | 25.545 | 4999 |
| 3725 | 1892 | 23.970 | 4911 | 2210 | 25.717 | 5032 |
| 3750 | 1896 | 23.999 | 4925 | 2225 | 25.890 | 5066 |
| 3775 | 1901 | 24.028 | 4938 | 2239 | 26.062 | 5100 |
| 3800 | 1905 | 24.057 | 4952 | 2254 | 26.235 | 5134 |
| 3825 | 1909 | 24.085 | 4965 | 2269 | 26.408 | 5168 |
| 3850 | 1913 | 24.113 | 4979 | 2284 | 26.580 | 5201 |
| 3875 | 1918 | 24.141 | 4992 | 2299 | 26.753 | 5235 |
| 3900 | 1922 | 24.169 | 5005 | 2314 | 26.925 | 5269 |
| 3925 | 1926 | 24.196 | 5019 | 2328 | 27.098 | 5303 |
| 3950 | 1930 | 24.224 | 5032 | 2343 | 27.271 | 5336 |
| 3975 | 1935 | 24.251 | 5045 | 2358 | 27.443 | 5370 |
| 4000 | 1939 | 24.278 | 5058 | 2373 | 27.616 | 5404 |
| 8000 | 3878 | 48.557 | 10117 | 4746 | 55.232 | 10808 |

MAD: Mean Amplitude Deviation, MIMS: Monitor Independent Movement Summary, ENMO: Euclidean Norm Minus One

**Figure S1: Exclusion flowchart.**

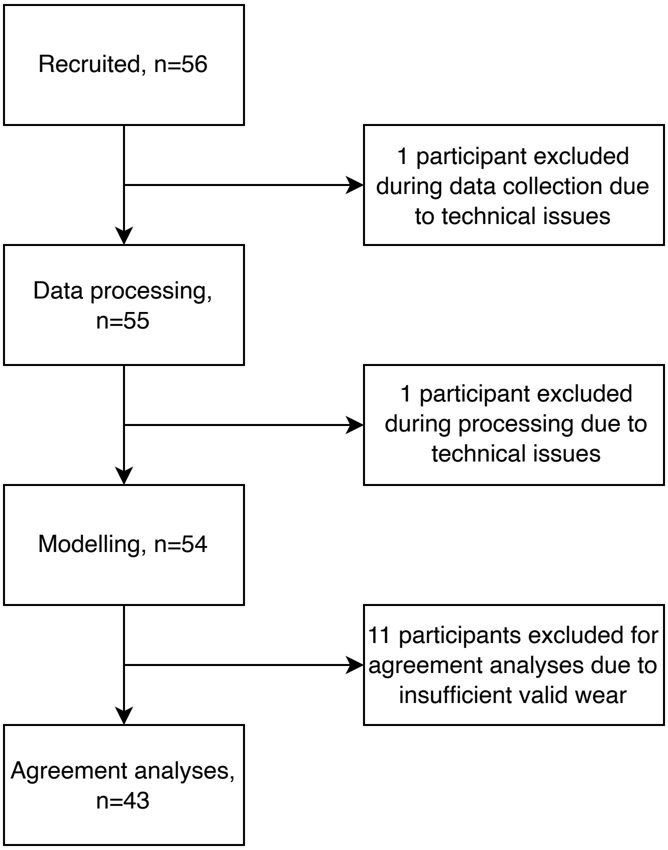
Recruited, n=56
1 participant excluded during data collection due to technical issues
Data processing, n=55
1 participant excluded during processing due to technical issues
Modelling, n=54
11 participants excluded for agreement analyses due to insufficient valid wear
Agreement analyses, n=43

**Figure S2: Bland-Altman plots showing bias and limits of agreement for intensity gradients generated based on naïve and modelled approaches for each of MAD, MIMS, and counts compared to ENMO with outlier excluded.** X-axes: means of paired IGs, y-axes: differences between paired IGs (each of MAD, MIMS, and counts minus ENMO). IG: Intensity gradient, MAD: Mean Amplitude Deviation, MIMS: Monitor Independent Movement Summary, ENMO: Euclidean Norm Minus One

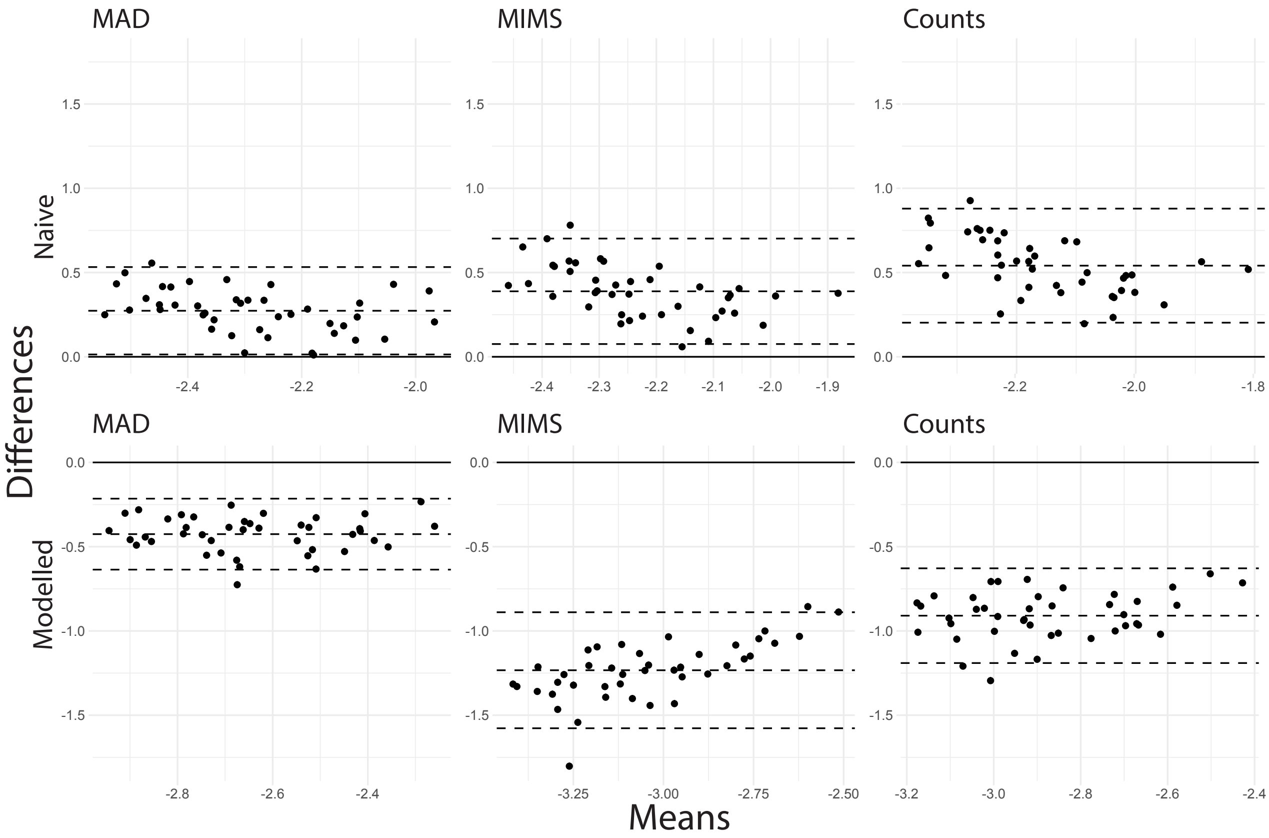

MAD
MIMS
Counts
Naive
Modelled
Differences
Means
